# The Epidemiologic Transition Theory and Evidence for Cancer Transitions in the US, Select European Nations, and Japan

**DOI:** 10.1101/2020.11.25.20238832

**Authors:** Omer Gersten, Magali Barbieri

## Abstract

Despite cancer being a leading cause of death worldwide, scant research has been carried out on the existence of “cancer transitions,” the idea that as nations develop, they move from a situation where infectious related cancers are prominent, to one where non-infectious related cancers dominate. We use annual cause-of-death data to produce death rates for common types of cancer in select high-income countries. We find that cancer mortality patterns parallel the epidemiologic transition, which states that as countries advance, they move from a regime where infectious diseases are most common to one where non-infectious disease are most common. An implication is that the epidemiologic transition theory as originally formulated continues to be relevant despite some researchers arguing that we need additional stages beyond the original three.

## Introduction

Cancer is one of the leading causes of death worldwide and because of marked declines in cardiovascular disease in several countries, cancer has become the leading cause of death in many high-income populations.^1,2,3^ While the important role of infection in causing certain cancer types is established and quantified (2.2 million cancer cases were attributable to infection in 2018), ^4,5,6^ the classical view of cancer as a prime example of a “degenerative and man-made disease”^7,8,9^ remains. Indeed, most global reports, including those of the World Health Organization (WHO), categorize cancer as a key non-communicable disease (NCD).^10^

Abdel Omran’s seminal article on the epidemiologic transition has had a deep and wide-ranging influence on a number of academic fields^11,12^ and has been cited nearly 5,000 times.^13^ In his article, Omran predicted that in the third and last stage of the transition, non-communicable and chronic ailments like heart disease and cancer replace infectious diseases as prime killers. In this age of degenerative and man-made diseases, infectious diseases experience a “progressive decline,” but do not disappear entirely.

Omran’s classical view, though, is overly simplified because it overlooks the fact that infection is often an important cause of cancer.^14^ For instance, we now know that the bacterium *Helicobacter pylori* (*H. pylori*), hepatitis B and C viruses, and human papillomavirus are important causes of cancers of the stomach, liver, and cervix, respectively. The number of new cancer cases in 2012 attributable to infectious agents was about 79% for stomach cancer, 73% for liver cancer, and 100% for cervix uteri cancer.^14^

Gersten and Wilmoth^15^ first introduced and developed the concept of the “cancer transition,” which they meant to be analogous to Omran’s epidemiologic transition. The authors analyzed cancer trends in Japan from 1951 until 1997 and found that cancers with a root in infectious causes were declining, while those with a root in non-infectious causes were increasing. There has thus been a playing out of the epidemiologic transition within the broad and complex set of diseases that define cancer.

Work investigating and attempting to extend the cancer transition theory has been exceptionally limited, perhaps numbering no more than a few papers. One book chapter, though, that has seriously engaged with the theory is that by Knaul and colleagues^16^ which at one point presents cross-sectional data from GLOBOCAN (2008) suggesting that cancers with a root in infectious causes become less prominent as many nations move along the gradient from less to more developed. A similar finding has been reported by Fidler and others^17^ in a more recent paper using a different dataset.

Bray and co-authors^4^ also extend the cancer transition literature. They analyze four levels of the Human Development Index – a composite indicator of life expectancy, education, and gross domestic product per head – to highlight cancer-specific patterns in 2008 and trends from 1998-2002, and to produce future burden scenarios for 2030. One of their conclusions is that “The reduction in infection-related cancers seems to be offset by concomitant increases in cancers related to a westernization of lifestyle and changes in tobacco consumption and the effect on lung and other cancers.” (page 800).

The aim of the paper here, then, is to contribute to the nascent literature on cancer transition theory, and in so doing, examine the relevance of Omran’s epidemiologic transition theory. The paper focuses on evidence of cancer transition in the US, select European nations, and Japan, differentiating between cancers with an infectious root and other cancers. We define the cancer transition as the mid-point at which a country’s population moves from a situation where rates of infectious-related cancers are greater than those of non-infectious cancers, to the converse situation where rates of non-infectious cancers are higher.

## Results

Figure 1 shows trends in the age-standardized death rates from infectious-related and non-infectious related cancers for all ages and both sexes combined in the six countries investigated here. In all countries, mortality from infectious-related cancers has declined steadily throughout the time period, except for Japan. At the beginning of the study period, the age-standardized death rates from cancers with a root in infectious causes ranged from about 50 per 100,000 in the US to 160 per 100,000 in Japan, a ratio of 1 to 3.2. Note, however, that the series starts in 1959 in the former country, but in 1950 for the latter one. In Japan, the only country where mortality from these cancers increased, the peak was reached at around 180 per 100,000 in 1955. At the end of the period (2016-2018, depending on the country), the rates range from about 20 per 100,000 in Norway, Sweden, England and Wales, and the US to nearly 60 per 100,000 in Japan. The discontinuity in the data is most visible for Japan and France, and is due to changes in the classification of liver cancer.

**Figure 1.**
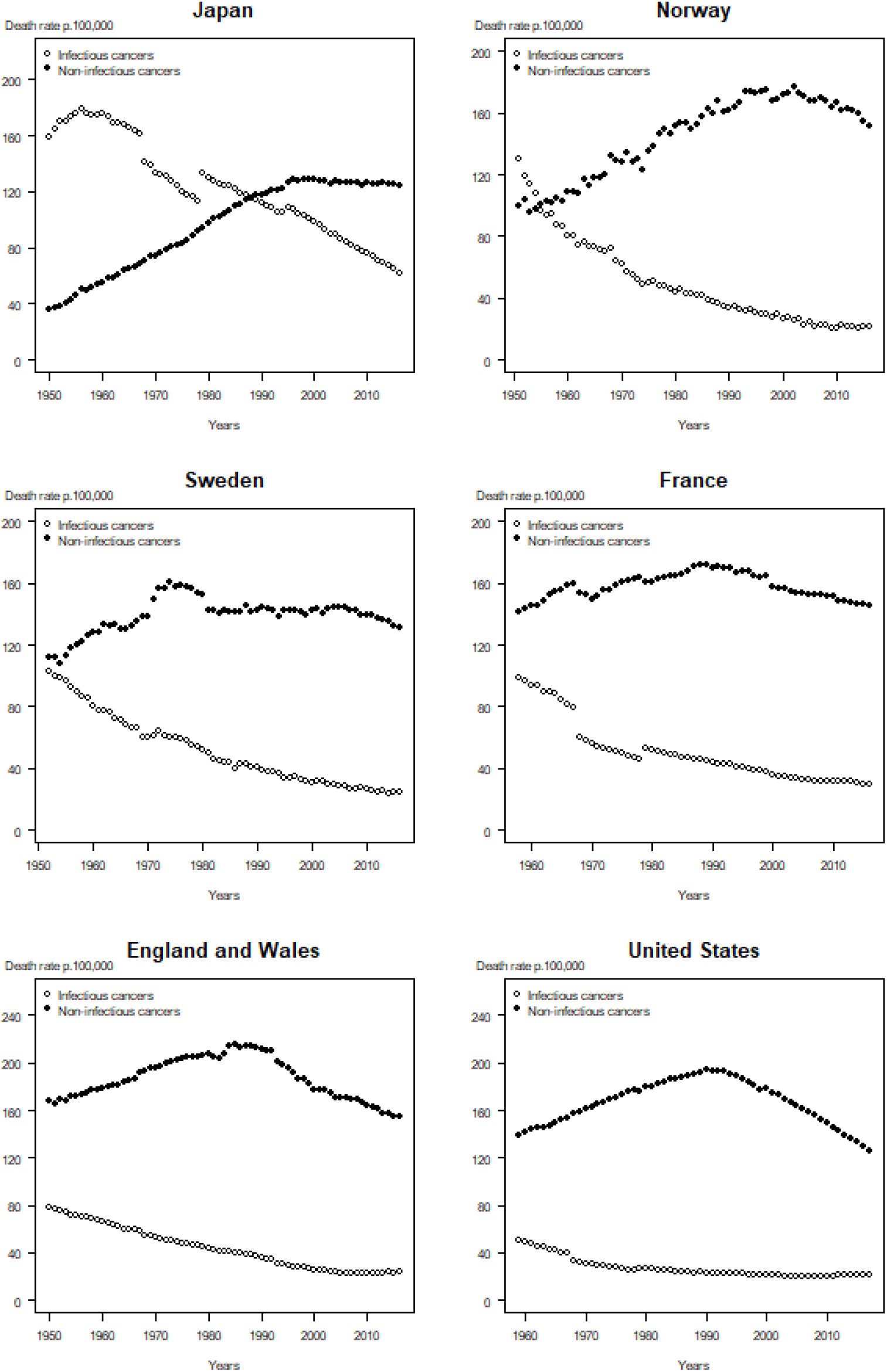
The cancer transition, both sexes combined, in Japan, select European nations, and the US.

At the beginning of the study period, all countries exhibited an increasing trend in the non-infectious related cancers, with a maximum reached circa 1985 in England and Wales. Most of the other countries experienced a peak around 1990, or a few years later in the case of Japan. In all but the US and England and Wales, mortality from non-infectious related cancers were at lowest at the beginning of the study period. Initial rates varied more than for infectious-related cancers, ranging from about 40 per 100,000 in Japan to about 170 per 100,000 in England and Wales, a ratio of 1 to 4.25. The peak also varied from one country to another, with a minimum of nearly 120 per 100,000 in Japan (from the mid-1990s to the end of the study period) up to over 200 per 100,000 in England and Wales (circa 1985). While the rates have remained high with little signs of decline in Japan, rates for other countries have begun to decline.

Japan and Norway are the only two countries where trends in mortality from the two groups of cancers intersect (in the late 1980s in Japan and in the mid-1950s in Norway). These countries completed their cancer transition latest in the time period; on the basis of back-extrapolation, it is likely that Sweden would have completed its transition a few years earlier than Norway, with France, England and Wales, and the US experiencing their transitions earlier than Sweden.

## Discussion

In this study we used temporal cancer-specific mortality data to assess whether there is supporting evidence of a cancer transition in the US, England and Wales, France, Norway, Sweden, and Japan. Overall, there is compelling evidence supporting the cancer transition theory.

Of the over one hundred cancers responsible for the total cancer burden, the nine cancers we examined account for over two-thirds of all cancer deaths worldwide in 20181. In investigating the extent of the transition from infectious to non-infectious-related cancers, we have identified specific time points when Japan and Norway had their transitions (when the trends converge) and speculated on the timing of the other nations studied. While all six countries selected are all classified as high-income, they are considerably diverse geographically, politically, and culturally, yet their cancer patterns are quite similar.

Visually, Japan exhibits the most obvious cancer transition in the countries we examined since its trends intersect later in the time period for which we have recorded data and most closely resembles an “X” pattern. Norway is the only other country where a cancer transition cross-over is visible, but it is easy to imagine that similar intersections would be observed elsewhere, were additional historical data available. If Japan might be considered a “laggard” in terms of its cancer transition, the US or England and Wales might be considered “forerunners,” since their transitions appear to have taken place in the first half of the twentieth century, prior to mortality data becoming available if one extrapolates back from visible trends.

We can go further in our description of cancer transitions by making some generalizations about their evolution: 1) initially, infectious-related cancer mortality rates are high and non-infectious-related cancer mortality are low; 2) then, infectious-related cancer mortality starts declining, while non-infectious cancer mortality increases, 3) subsequently, infectious-related cancer mortality reaches a low level while non-infectious cancer mortality starts declining after reaching its peak.

Of primary importance in our paper was the grouping and analysis of infectious-related versus non-infectious related cancers to identify signs of cancer transition. A caveat to such an analysis is the extent to which the overall trends mask some of the underlying diversity in individual cancers by sex (see Supplementary Figures 1-2). Still, despite concealing individual heterogeneity, grouping cancers with more or less infectious-orientated etiologies, shows a concise picture of cancer transitions.

To conceptualize the cancer transition that occurs as nations develop, we apply elements of Omran’s theory of the epidemiologic transition.^7^ Omran proposed that there are three stages in the epidemiologic transition, the last being a move to the “age of degenerative and man-made diseases” where illnesses like stroke and cancer rise in prominence as infectious diseases like malaria, tuberculosis, and smallpox become less common. We consider Omran’s understanding at the time of cancer as a non-communicable/non-infectious-related disease the “classical view” of cancer’s etiology. In the last stage explicated by Omran, infectious diseases are increasingly brought under control, but not entirely. Historically, infectious diseases tended to strike at younger ages, and when these diseases were better brought under control people lived to increasingly older ages when non-communicable diseases tended to strike.

Some researchers have attempted to extend Omran’s three-stage theory by proposing a “fourth stage” or even “fifth stage” of the epidemiologic transition.^8,36,37^ For instance, Olshansky and Ault^37^ postulated that there’s a fourth stage of the transition that they term the “age of delayed degenerative diseases” in which major causes of death are still with us, but the risk of dying from those illnesses is redistributed to older ages. Like many others during their time and continuing to this day,^8-10^ Olshansky and Ault adopt the classical view of cancer as a non-communicable disease.

Emerging mainly from the anthropological literature is the idea that we are in a “third transition,” not to be confused with the “stages” of Omran’s epidemiologic transition.^38,39^ According to this literature, the first transition is one that predates the first stage described by Omran, and occurred in the Neolithic period about 10,000 years ago. Authors writing on this topic also state that at about this time there was a shift marked by the emergence of many infectious diseases. The whole of Omran’s epidemiologic transition is considered to be the second transition, and in the third transition there are emerging infectious diseases (e.g., HIV) and a reemergence of infectious diseases (e.g., tuberculosis), many of which are multiple antibiotic resistant.

Even with emerging diseases like covid-19, deaths due to cancer, in the US at least, are far greater than those caused by infectious diseases. At the time of the writing of this paper, covid-19, which was first identified in December 2019, has so far killed about 250,000 Americans,^40^ but scientists have developed a vaccine. Influenza and pneumonia, which kill people year after year and are also infectious, caused about 55,000 deaths in the US in 2017.^41^ In contrast, about 600,000 people died of cancer in that same year and country^41^, and deaths due to infectious sources, for the US from 1980 to 2014, only accounted for a small portion (5.4%) of total deaths^42^. These figures, along with the fact that many so-called “new” infectious diseases are actually hundreds if not thousands of years old,^43^ argue against a new stage of the epidemiologic transition based on emerging and re-emerging infectious diseases.

While some proposed extensions of Omran’s epidemiologic transition have made useful contributions to the literature, they do not grapple with the cancer transition theory as we have laid out in this paper. What many authors have not realized is that a number of widespread cancers are mainly caused by infectious agents. To wit, *H. pylori* is a cause of stomach cancer, the human papillomavirus is a necessary, albeit insufficient, cause of cervix uteri cancer, and hepatitis B and C viruses are causes of liver cancer.^14^

As regards *H. pylori*, over half of the world’s population in 2015 were infected with the bacterium^44^, but only a small fraction of those infected will go on to develop stomach cancer^14^. *H. pylori’s* prevalence is declining in most countries of the world^45,46^ not through any specific measures, but as a result of general improvements in sanitation, housing conditions, access to clean water, food availability and freshness, and reduced salt intake.^15,47,48^

The human papillomavirus is transmitted sexually, with the majority of sexually active individuals of both sexes acquiring it at some point during their lifetime.^49^ Prior to the vaccine becoming available in 2006,^50,51^ screening and treatment were the only means to reduce cervical cancer incidence and mortality.^52^ Research is continuing into creating more affordable vaccines, which will especially help less developed countries where current levels of vaccination are a fraction of what they are in more developed countries.^51,53^ Reductions of cervical cancer will only be evident decades after vaccination, but a surrogate for cervical cancer – high-grade cervical intra-epithelial neoplasia – has shown significant reductions in populations with high vaccine coverage.^54^

The last cancer with a root in infectious causes that we investigated is liver cancer, and persistent hepatitis B and C viruses together account for about 90% of hepatocellular carcinoma, the most common form of liver cancer.^55^ A vaccine for hepatitis B has been available since the early-1980s, and was progressively administered to infants as part of national childhood immunization programs in 90 countries in the late 1980s and 1990s. Hepatitis B decreases have been observed in most countries.^56,57^ Currently there is not a vaccine for hepatitis C, but there is antiviral treatment which cures about 90% of those infected with the virus.^58^ To summarize, there are reasons to expect that mortality from cancers with a root in infectious causes will decline even further in the future.

As far as the trends for the combined non-infectious related cancers are concerned, there was a plateauing or decline in five of the six countries starting between 1985-2000 (with Sweden’s decline occurring in the early 1970s). The pattern of risk factors over the last few decades in these countries for the individual cancers are mixed. A decline in total physical activity has been documented in Japan^59,60^ and the US.^61^ Although there doesn’t appear to be country-specific data for the other nations we examine, one study found that in high-income Western countries, between 2001 and 2016, physical inactivity increased by 5%.^62^ Another important trend is that Japan and France seem to have largely resisted “Westernization”/degradation of their diets,^63,64^ but not the US^65^ and England and Wales^66^. There is not enough literature on the diets in Norway and Sweden to determine the extent to which their diets have changed.

The average body mass index (BMI) in 2014 for Japanese men and women was in the “normal” range, but for the other countries, either men or women, or both, have average BMIs in the “overweight” category.^67^ Alcohol consumption from 1990 - 2016 for both men and women was nearly level in the US and Japan, slightly increased in England and Wales, Norway, and Sweden, and declined markedly in France.^68^ A substantial, positive trend, has been a reduction in smoking prevalence that has resulted in lower lung cancer rates some thirty to forty years later.^69^

Importantly, the ability to both detect and treat some cancers has improved dramatically, such as for cancers of the breast^70^ and colorectum.^71^ Prostate-specific antigen (PSa) screening remains controversial,^72,73^ but treatment for prostate cancer has become more effective.^74^ Unfortunately, esophageal cancer is typically diagnosed late in its course when it’s more difficult to treat, but in cases where early esophageal cancer is detected, evolving therapies have improved the cure rate.^75^ For pancreatic cancer, even though there has been advances in imaging and surgery, mortality rates have decreased little as a result.^76^

Based on our findings, we confirm and extend the hypothesis of Gersten and Wilmoth^15^ that the cancer transition can be considered analogous to the epidemiologic transition. Put another way, one can say that the cancer transition “parallels” the epidemiologic transition, in that there is an epidemiologic transition also within the complex system of cancers linked to specific causes. This linkage between the epidemiologic and cancer transition theories is presented schematically in Figure 2.

**Figure 2.**
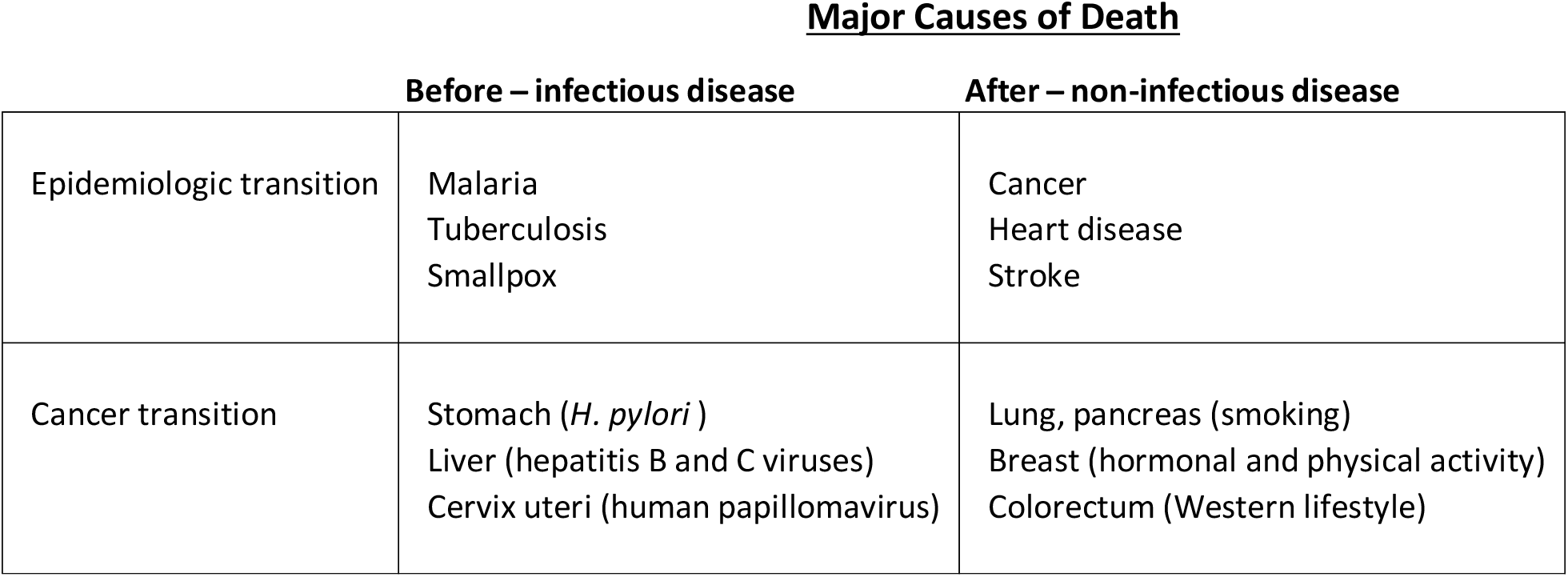
Comparison of the epidemiologic transition and cancer transition theories.

A main implication of our analysis, then, is that while Omran’s theory does not apply to part of the third stage of cancer transitions, in which non-infectious related cancers have peaked and have in some settings, begun to decline, cancer remains bound in epidemiologic transition theory. Indeed, the theory continues to remain a very useful framework for understanding fundamental mortality patterns for cancer, a leading cause of death worldwide.

## Methods

We selected the countries in our analysis based on the availability, detail and quality of their cause-of-death records, and how far back the data series extended. Annual cause-of-death data were obtained directly from national statistics offices and the World Health Organization (WHO) Mortality Database^18^ and combined with life table series from the Human Mortality Database^19^ to compute age-standardized death rates for common types of cancer in six countries over extended periods of time: the United States (1959-2017), England and Wales (1950-2018), France (1958-2016), Norway (1951-2016), Sweden (1952-2016), and Japan (1950-2016). The European 2013 standard population was used to account for differences in age structure across the six countries.

Our analysis presents results for all ages combined and for nine cancers grouped according to their known etiology, separating those with a root in infectious causes from those with non-infectious causes. Cancers with a root in infectious causes included stomach, liver, and cervical cancer, which together accounted for 19.7% of worldwide cancer deaths in 2018^1^

Concerning the six cancers with a root in non-infectious causes that we analyze, those cancers and their main causes are: lung – smoking;^20^ pancreatic – smoking;^21,22^ esophageal – smoking and drinking;^23,24^ breast – reproductive/hormonal and physical activity;^25,26^ colorectal – Western lifestyle;^27,28^ and prostate – age, ethnicity, and family history.^29-31^ Together, these cancers accounted for 47.6% of worldwide cancer deaths in 2018.^1^

The International Classification of Diseases (ICD), implemented by all study countries during the time period under consideration, has gone through numerous revisions, from the sixth in 1950 to the tenth currently. These ICD revisions introduce some disruptions in the data series. However, as documented in the literature, cancer as a disease is much better defined than many others and these disruptions are small enough that a general picture of the overall trends emerges very clearly.^32,33^ Similarly, differences in diagnostic and coding practices do not appear to be large enough across countries to bias international comparisons in cancer mortality trends.^34,35^

## Data Availability

All the data can be accessed for free at the Human Mortality Database

https://www.mortality.org/

## Acknowledgements

We are grateful to Freddie Bray and Ken Wachter for their comments on a draft of this paper, and we wish to thank Yuan Zhang for organizing a synchronous, virtual session for the 2020 Population Association of America Annual Meeting at which this paper was presented orally. Lastly, we thank Miriam Rauch for providing vital support.

---

**Supplementary Figure 1.**
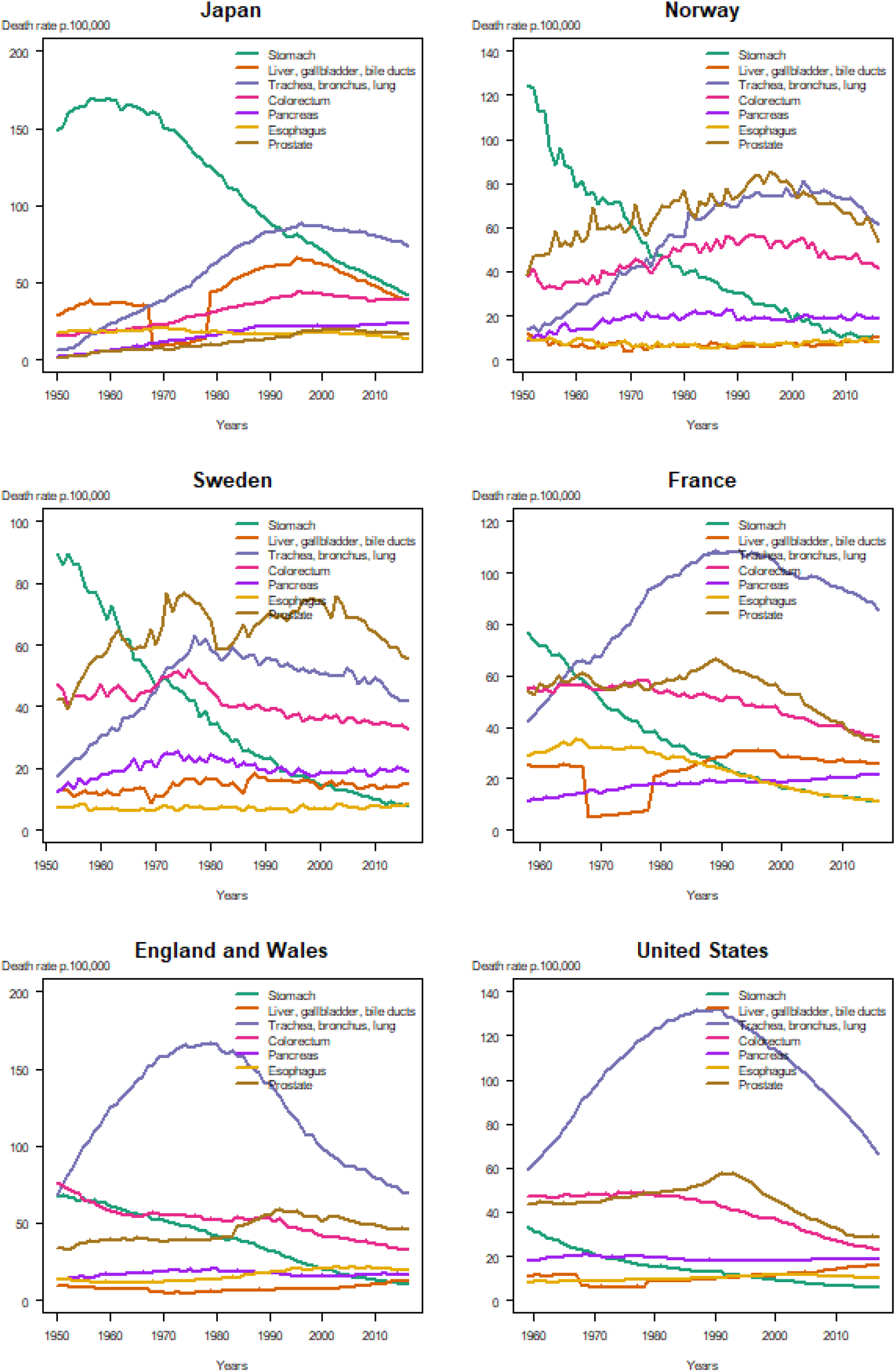
Age-standardized death rates for the most common cancers over time, men, in Japan, select European nations, and the US.

**Supplementary Figure 2.**
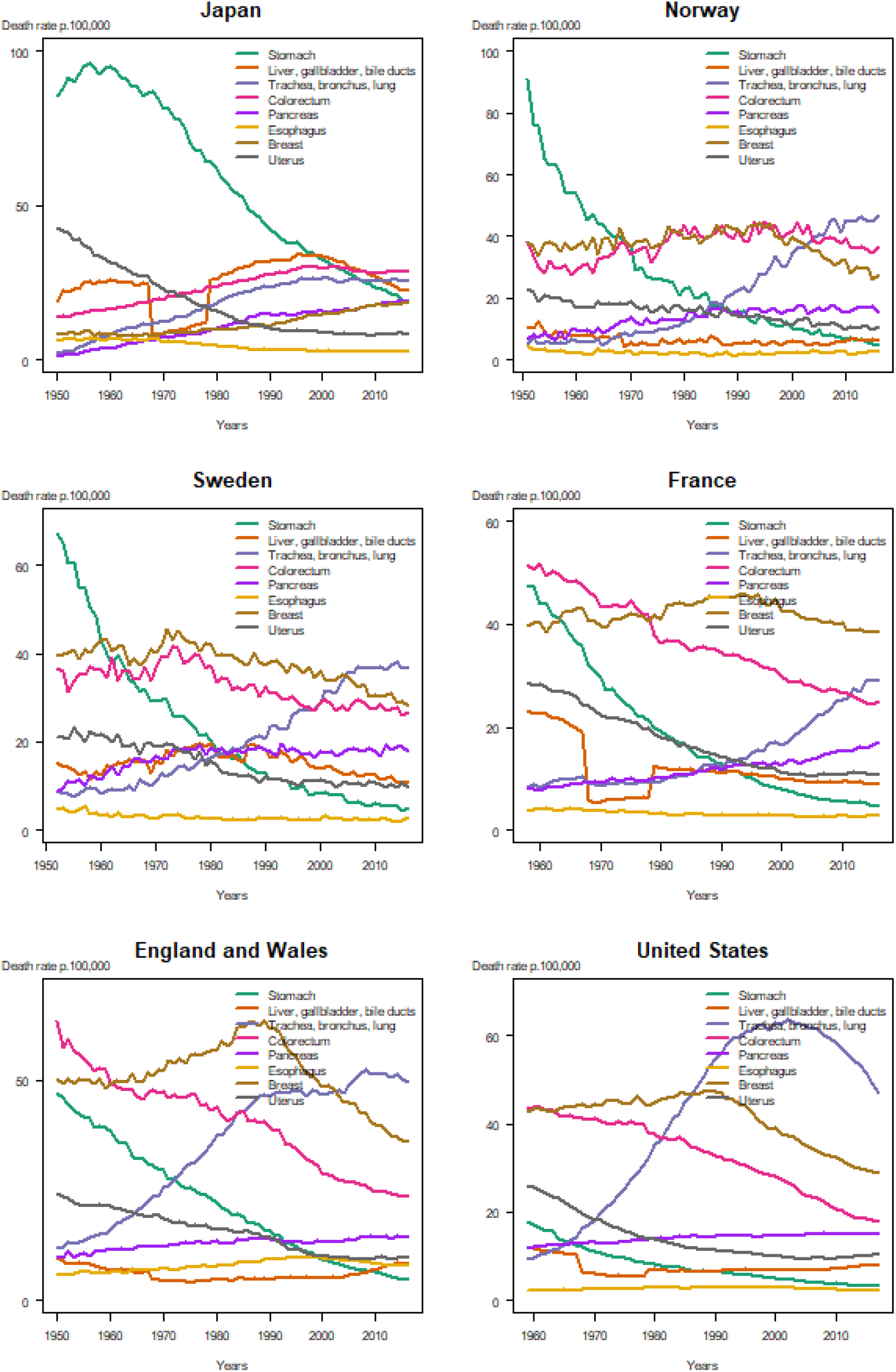
Age-standardized death rates for the most common cancers over time, women, in Japan, select European nations, and the US.

